# Effects of CPAP on Atherosclerotic Coronary Plaques in Patients with Sleep-Disordered Breathing: the ENTERPRISE Trial

**DOI:** 10.1101/2023.08.31.23294929

**Authors:** Tatsuya Fukase, Tomotaka Dohi, Takatoshi Kasai, Naotake Yanagisawa, Ryota Nishio, Mitsuhiro Takeuchi, Norihito Takahashi, Hirohisa Endo, Hideki Wada, Shinichiro Doi, Hiroki Nishiyama, Iwao Okai, Hiroshi Iwata, Seiji Koga, Shinya Okazaki, Katsumi Miyauchi, Hiroyuki Daida, Tohru Minamino

## Abstract

**Background:** The effects of continuous positive airway pressure (CPAP) treatment on coronary atheroma volume and vulnerability in patients with sleep-disordered breathing (SDB) coexisting with coronary artery disease are not well known. Thus, this study aimed to verify how CPAP treatment affects coronary plaques in patients with SDB who underwent percutaneous coronary intervention (PCI).

**Methods:** Altogether, fifty-three patients with SDB who underwent elective PCI using near-infrared spectroscopy (NIRS) and intravascular ultrasound (IVUS) were randomly assigned to two groups based on CPAP use. This study evaluated the absolute changes in percentage atheroma volume (PAV) and total atheroma volume by IVUS, and the absolute changes in maximum lipid core burden index calculated for every 4-mm segment (maxLCBI_4mm_) using NIRS from baseline to 12 months, in addition to major adverse cardiac and cerebrovascular events (MACCE), and all-cause death.

**Results:** In the final analysis, 23 patients were allocated to the CPAP group, and 24 patients were allocated to the non-CPAP group. Compared with the non-CPAP group, the CPAP group had a significantly higher absolute change in PAV (−1.90±2.63% vs. 0.82±4.67%, *p*=0.018) and TAV (−20±35 mm^3^ vs. −2±21 mm^3^, *p*=0.041), and lower incidence rate of MACCE, mainly driven by a decrease in target lesion or non-target lesion target vessel revascularization (4% vs. 25%, *p*=0.037). In addition, adequate CPAP treatment resulted in greater plaque regression of −2.48%. However, there was no significant difference for absolute change in maxLCBI_4mm_ (0 [−32, 40] vs. 34 [−71, 163], *p*=0.306) between the both groups, and no all-cause death was observed.

**Conclusions:** CPAP treatment for patients with SDB contributed to a decline in coronary plaque volume and prevention of revascularization risk but did not mitigate lipid-rich plaques. Especially, adequate CPAP treatment had a greater effect on coronary plaque regression.

**Clinical Trial Registration:** The Ethics Committee of the Juntendo Clinical Research and Trial Center approved this study (reference number 18-021), and which was registered with the UMIN (UMIN ID: R000036293). https://upload.umin.ac.jp/cgi-open-bin/icdr_e/ctr_view.cgi?recptno=R000036293

**Clinical Perspective:** The effects of continuous positive airway pressure (CPAP) treatment on coronary atheroma volume and vulnerability in patients with sleep-disordered breathing (SDB) coexisting with coronary artery disease remains unclear. This study revealed that CPAP treatment for patients with SDB who underwent percutaneous coronary intervention contributed to a decline in coronary plaque volume and the prevention of cardiovascular events, mainly driven by a decrease in target vessel revascularization. In particular, adequate CPAP treatment may have a greater effect on coronary plaque regression; thus, it is important to focus on CPAP treatment and subsequent treatment compliance considering coronary plaques and events. In addition, patients with acute coronary syndrome who have greater plaque volume and lipid-rich plaques than those with stable coronary artery disease may benefit from the effects of adding CPAP treatment to optimal medical therapy due to an anti-atherogenic potential effect of CPAP treatment, thus, the further investigations targeting acute coronary syndrome are necessary.

## Introduction

Sleep-disordered breathing (SDB) is an important risk factor for coronary artery disease, and is strongly associated with hypertension, coronary artery disease, heart failure, arrhythmia, stroke, and cardiovascular mortality.^1, 2^ Especially in coronary artery disease, SDB has been associated with a 2-fold increased risk of cardiovascular events and death.^3, 4^ In addition, the degree of SDB is involved in the quantity and quality of coronary plaques detected by coronary computed tomography angiography.^5^ A report on the culprit plaque characteristics identified by intravascular ultrasound (IVUS) in patients who diagnosed with SDB using nocturnal pulse oximeter and underwent percutaneous coronary intervention (PCI) showed that SDB was associated with a larger atheroma plaque volume and greater ultrasound attenuation.^6^ Thus, SDB treatment is thought to lead to the regression of atherosclerosis, and continuous positive airway pressure (CPAP), an established SDB treatment, may be useful for coronary plaque regression and stability.^7^ However, there have been no reports on the effects of CPAP on coronary atheroma volume and vulnerability in patients with SDB and coexisting coronary artery disease. Thus, the effects of CPAP on atherosclerotic coronary plaques in patients with SDB and coronary artery disease (ENTERPRISE) trial aimed to evaluate the effects of adding CPAP treatment to optimal medical treatment on coronary plaques in patients with SDB who underwent PCI, using near-infrared spectroscopy (NIRS)-IVUS, which can simultaneously evaluate the plaque morphology and presence of lipid-rich plaques and is useful for identifying vulnerable plaques.^8, 9^

## Methods

### Study design, study population and data collection

The ENTERPRISE study was a prospective, randomized, 12-month, open-label, single-center study. The detailed protocol of the ENTERPRISE study has been described previously.^10^ Inclusion criteria for this study were stable coronary artery disease, successful PCI under IVUS guidance to treat a culprit lesion and evaluate a non-culprit lesion segment by NIRS-IVUS, and SDB defined as 3% oxygen desaturation index >15 events/h, determined using an overnight pulse oximeter. Patients were prospectively enrolled at Juntendo University Hospital from July 2018 to July 2021. The exclusion criteria were current or previous CPAP treatment, hypersomnia requiring urgent treatment defined as an Epworth Sleepiness Scale score ≥18, New York Heart Association class II, III, or IV heart failure, renal insufficiency (serum creatinine ≥2.0 mg/dL) and hemodialysis, active malignant disease, and patients recognized as unsuitable by the attending physician.

The study protocol was approved by the ethics committee (PR052/08, 07/064/797). The Ethics Committee of the Juntendo Clinical Research and Trial Center approved this study (reference number 18-021), which was registered with the University Hospital Medical Information Network (ID: R000036293). All the participants provided written informed consent. The study conformed to the principles outlined in the Declaration of Helsinki.^11^

Eligible patients were randomized to receive individualized lifestyle counseling therapy plus CPAP (CPAP group) or conservative treatment (non-CPAP group) consisting only of individualized lifestyle counseling therapy. The randomization was stratified using permuted block randomization with two stratification factors; body mass index <25 kg/m^2^ or ≥25 kg/m^2^, and 3% oxygen desaturation index <30 events/hour or ≥30 events/hour.

Demographic data, coronary risk factors, and medication information were obtained from the institutional database. Blood samples were collected before the procedure. Based on the baseline low-density lipoprotein cholesterol (LDL-C) level, attending doctors selected lipid-lowering agents as secondary prevention to meet the target LDL-C level of <70 mg/dL, in addition to diet and exercise therapy, as recommended by the latest guidelines.^12^ Thus, the statin dose was increased until the target LDL-C level was reached, and if the target LDL-C level was not reached after administration of high-dose statins, lipid-lowering agents were administered. The high doses of atorvastatin, pitavastatin, and rosuvastatin were 20 mg, 4 mg, and 10 mg, respectively. Also, some patients received treatment with anti-hypertensive agents if they had a blood pressure >140/90 mmHg, glycemic control and management of comorbidities from diabetes if they had a hemoglobin A1c ≥6.5 % or fasting plasma glucose ≥126 mg/dL, and smoking cessation guidance. Safety was monitored throughout the study and evaluated by periodic medical examination and laboratory tests every two months after enrollment.

### Sleep studies

The presence of SDB was determined using a wristwatch-type pulse oximeter (PULSOX-Me300; Konica Minolta, Tokyo, Japan). Overnight pulse oximetry was performed during hospitalization before the elective PCI. The value of the 3% oxygen desaturation index, which was defined as the index of oxygen desaturation represented as the number of events per hour of the recording time in which the patient’s blood oxygen level fell by ≥3 %, was used as an indicator of SDB severity. In this study, SDB was defined as a 3% oxygen desaturation index ≥15 events/hour, and severe SDB was defined as a 3% oxygen desaturation index ≥30 events/hour, based on a previous report.^13^ Also, this device recorded the average and minimum saturation of percutaneous oxygen, total measurement time for pulse oximetry, and cumulative time with saturation of percutaneous oxygen <90 %.

The Epworth Sleepiness Scale was used to investigate daytime sleepiness and contained eight questions to evaluate the chance of dozing off under eight scenarios in the past month. Each item was scored from 0 to 3 (0=never doze; 1=slight chance of dozing; 2=moderate chance of dozing; 3=high chance of dozing), and Epworth Sleepiness Scale score ranged from 0 to 24. Excessive daytime sleepiness was defined as an Epworth Sleepiness Scale score ≥10.^14^ All patients were scored at baseline and follow-up.

### NIRS-IVUS imaging acquisition and analysis

Non-culprit lesions were defined as any lesions in the entire coronary artery outside the culprit lesion and were imaged using NIRS-IVUS pullback at both baseline and 12-month follow-up. NIRS-IVUS was performed using a commercially available system (TVC Imaging System or Makoto Imaging System; Infraredx, Bedford, MA, USA). This modality combines the functions of grayscale IVUS and NIRS, which identify the chemical components of coronary artery plaques as a means of assessing vulnerability.^15^ A NIRS-IVUS catheter was inserted distal to the non-culprit lesion and pulled back at a rate of 0.5 mm/sec, after intracoronary injection of nitroglycerin.

The NIRS-IVUS analysis was accurately performed by two cardiologists (T.F. and T.D.), who were blinded to the sleep information obtained from pulse oximeter and the intervention of CPAP. The NIRS images were analyzed off-line, and the following parameters were measured. The NIRS chemogram presented data in yellow indicating the presence of lipid-rich plaque, or red indicating the absence of lipid-rich plaque, and allowed calculation of the lipid core burden index (LCBI): total yellow pixels divided by total viable pixels within the region of interest multiplied by 1000.^16^ In this study, the LCBI was calculated for every 4-mm or 10-mm segment within the non-culprit lesion, and these maximum values were defined as the maximum LCBI within any 4-mm long segment (maxLCBI_4mm_) and 10-mm long segment (maxLCBI_10mm_), respectively. A large lipid-rich plaque was defined as lipid-rich plaque with a maxLCBI_4mm_ ≥400, which was the cutoff value validated in previous studies.^17, 18^

Quantitative grayscale IVUS measurements were performed using QIvus version 2.1 (Medis, Leiden, the Netherlands) to quantify lumen cross-sectional area, external elastic membrane cross-sectional area, plaque and media cross-sectional area, and plaque burden, according to two clinical expert consensus documents.^19, 20^ In addition, this software can measure vessel volume, total atheroma volume and percentage atheroma volume. Regarding plaque morphology, atheroma is classified as follows: fibrous plaque has an intermediate echogenicity between soft (hypoechoic) plaque and highly echogenic calcified plaques, and calcified plaque has a higher echogenicity than the adventitia with an acoustic shadow.^19, 20^

### CPAP intervention

For patients randomized to receive CPAP treatment, an auto-CPAP machine (Sleepmate 10 Auto; ResMed, Sydney, Australia) was used to measure and store information about respiratory events (apneas and hypopneas), patient use (minutes used and nights used), mask leaks, and pressure, delivered using different masks ranging from small nasal pillows to a full-face mask, depending on patient preference. The attending physician confirmed the usage status of CPAP and provided guidance on mask fitting and CPAP pressure adjustment at least once per month. In addition, a system was established to enable communication with patients as needed, and prompt responses were provided if any issues arose. Adequate CPAP treatment was defined as both ≥4 hours daily use and ≥75% monthly use.^21^

### Endpoints

The primary endpoint was defined as the absolute change in the coronary atheroma volume of the non-culprit lesion measured using IVUS from baseline to 12 months, calculated as follows: percentage atheroma volume (PAV) = ∑ (external elastic membrane cross-sectional area − lumen cross-sectional area) / ∑ external elastic membrane cross-sectional area × 100.^20^

This study defined the secondary endpoints as: absolute changes in the extent of lipid-rich plaque in the non-culprit lesion by NIRS analysis; absolute change in total atheroma volume of the non-culprit lesion by IVUS analysis; absolute change in maximum intima media thickness by carotid ultrasonography; absolute change in serum high-sensitivity C-reactive protein level; major adverse cardiac and cerebrovascular events (MACCE), defined as composite of cardiovascular death, non-fatal myocardial infarction, non-fatal stroke, hospitalization for unstable angina, target vessel revascularization including target lesion revascularization and non-target lesion target vessel revascularization, and hospitalization for congestive heart failure; and all-cause death.

### Statistical analysis

Categorical data are presented as number (percentage) and were compared using the chi-square test. Continuous variables were expressed as mean±standard deviation or as median (interquartile range [IQR]) and compared using Student’s t-test or Mann-Whitney U test. The Shapiro–Wilk test was used to examine whether the scores were likely to follow a certain distribution in all patients. If *p*<0.05, the variables were not considered normally distributed. The change in Epworth Sleepiness Scale score from baseline to follow-up was compared using a paired *t*-test in the CPAP group and non-CPAP group, respectively. The absolute changes in PAV, maxLCBI_4mm_, maxLCBI_10mm_, total atheroma volume, maximum intima-media thickness, and high-sensitivity C-reactive protein levels were evaluated using analysis of covariance, including baseline measurement as a covariate. Univariable analysis of the relationship between the PAV and maxLCBI_4mm_ or maxLCBI_10mm_ was performed using the Pearson correlation analysis. All probabilities were expressed as two-tailed values, with statistical significance set at *p*<0.05. All data were analyzed using JMP^®^ Pro, version 16.0.0 for Macintosh (SAS Institute, Cary, NC, USA).

The sample size of ENTERPRISE trial was calculated in a previously reported protocol.^10^ The target sample size had been set at 100 patients (50 patients per group), and this sample size was selected based on the number of patients deemed capable of being included within the duration of an exploratory clinical study. The standard deviation for percentage change in atheroma volume in the PRECISE-IVUS trial was 5.55%.^22^ In detail, the sample size of 100 patients was expected to be statistically significant if detecting a 2.20% difference in the primary endpoint, assuming a standard deviation of 5.55% with 5% type I error for a 2-sided 2-sample *t*-test.

## Results

### Patient’s clinical characteristics

A total of 209 patients were initially assessed of whom 156 were excluded. Finally, 53 patients were recruited, 26 of whom were assigned to the CPAP group and 27 to the non-CPAP group. Three patients in the CPAP group and three in the non-CPAP group discontinued follow-up; therefore, 47 patients completed the study and were analyzed. (Figure 1).

**Figure 1.**
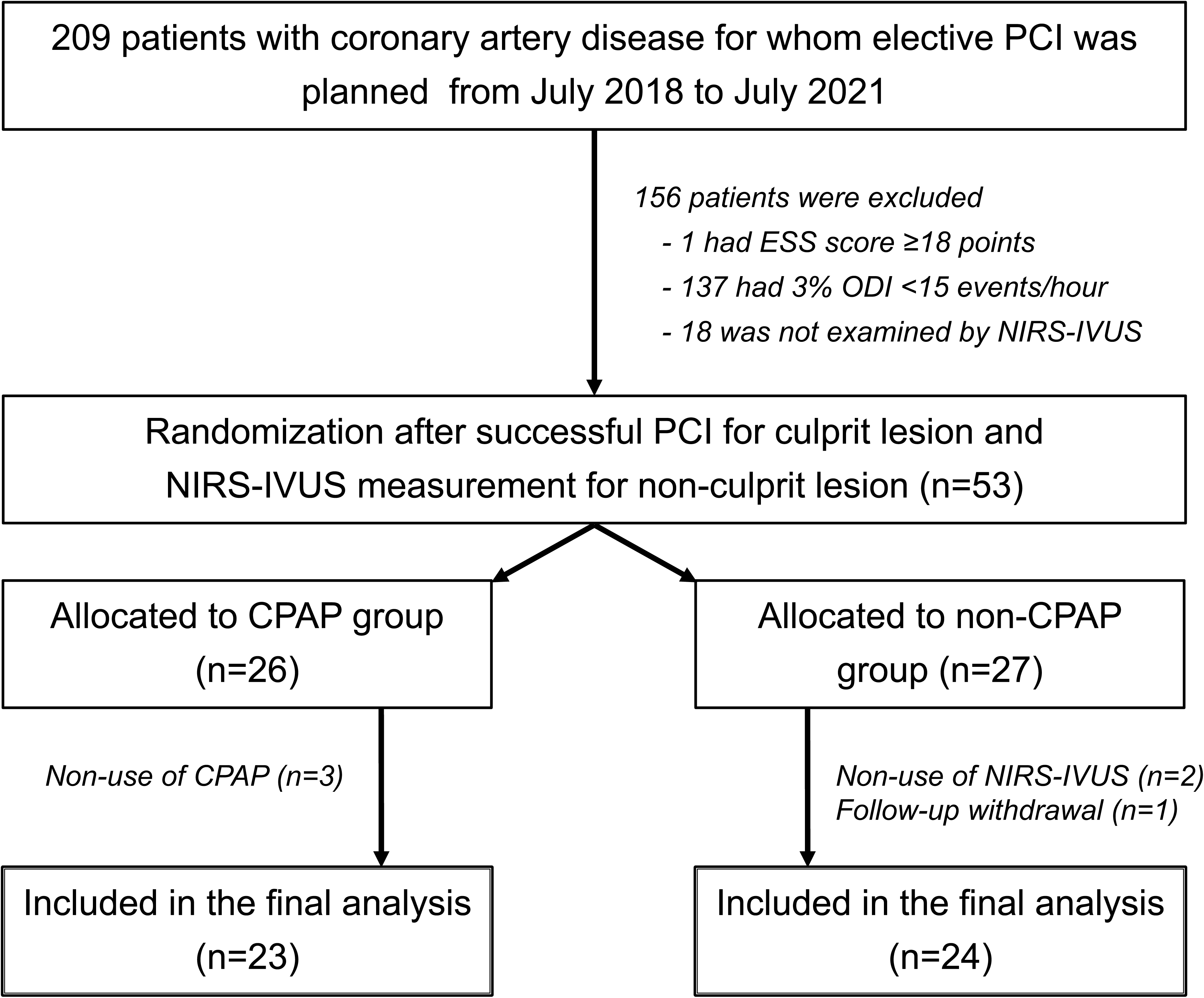
Flow chart. Among 209 patients with coronary artery disease for whom elective PCI was planned, 156 patients were excluded due to Epworth Sleepiness Scale score ≥18 points, 3% oxygen desaturation index <15, and non-usage of NIRS-IVUS. Finally, 53 patients with SDB who underwent successful PCI for culprit lesions and NIRS-IVUS for non-culprit lesions were recruited, 26 of whom were assigned to the CPAP group and 27 to the non-CPAP group. Because three patients in the CPAP group and three in the non-CPAP group discontinued the study at follow-up, 47 patients completed the study and were analyzed. CPAP, continuous positive airway pressure; NIRS-IVUS, near-infrared spectroscopy and intravascular ultrasound; PCI, percutaneous coronary intervention; SDB, sleep-disordered breathing.

The baseline clinical characteristics of the patients are summarized in Table 1. The mean patient age was 62±8 years, and 96% of them were males. The prevalence of hypertension, diabetes mellitus, current smoking, family history of premature coronary artery disease, chronic kidney disease, and history of myocardial infarction were 94%, 64%, 21%, 13%, 19%, and 40%, respectively. The baseline clinical characteristics of the patients were similar between the CPAP and non-CPAP groups, and there was no significant difference between the groups in terms of LDL-C levels, hemoglobin A1c levels, insulin resistance and proportion of statin use at both baseline and follow-up. Regarding the sleep parameters, baseline mean 3% oxygen desaturation index, proportion of severe SDB, average SpO_2_, minimum SpO_2_, total measurement time of pulse oximetry, and cumulative time with SpO_2_ <90 % were similar in the two groups.

**Table 1.** Patients’ clinical characteristics.

|  | Overall<br>(n=47) | CPAP<br>group (n=23) | Non-CPAP<br>group (n=24) | *p* value |
| --- | --- | --- | --- | --- |
| **Baseline characteristics** |  |  |  |  |
| Age, years | 62±8 | 62±8 | 62±8 | 0.946 |
| Male, n (%) | 45 (96) | 21 (91) | 24 (100) | 0.086 |
| Body mass index, kg/m<sup>2</sup> | 27.5±4.1 | 28.2±5.0 | 26.8±3.0 | 0.264 |
| Hypertension, n (%) | 44 (94) | 22 (96) | 22 (92) | 0.573 |
| Diabetes mellitus, n (%) | 30 (64) | 16 (70) | 14 (58) | 0.422 |
| Current smoker, n (%) | 10 (21) | 6 (25) | 4 (17) | 0.429 |
| Family history of premature CAD, n (%) | 6 (13) | 3 (13) | 3 (13) | 0.956 |
| Chronic kidney disease, n (%) | 9 (19) | 3 (13) | 6 (25) | 0.294 |
| History of myocardial infarction, n (%) | 19 (40) | 7 (30) | 12 (50) | 0.170 |
| **Laboratory data** |  |  |  |  |
| Low-density lipoprotein cholesterol, mg/dL | 78±23 | 80±19 | 77±26 | 0.658 |
| High-density lipoprotein cholesterol, mg/dL | 41±10 | 40±8 | 42±11 | 0.568 |
| Triglyceride, mg/dL | 159±97 | 168±96 | 151±99 | 0.567 |
| Lipoprotein (a), mg/dL | 17 (9, 40) | 18 (9, 30) | 16.5 (11, 52) | 0.663 |
| Apolipoprotein A-I, mg/dL | 119±22 | 116±19 | 121±25 | 0.442 |
| Apolipoprotein B, mg/dL | 79±20 | 81±16 | 78±23 | 0.568 |
| Hs-CRP, mg/dL | 0.07<br>(0.04, 0.45) | 0.07<br>(0.03, 0.59) | 0.08<br>(0.05, 0.36) | 0.949 |
| **Echographic data** |  |  |  |  |
| Left ventricular ejection fraction, % | 62±12 | 61±12 | 62±12 | 0.915 |
| Right maximum IMT, mm | 1.0 (0.9, 1.2) | 1.0 (0.9, 1.3) | 1.0 (1.0, 1.2) | 0.625 |
| Left maximum IMT, mm | 1.0 (0.9, 1.3) | 1.0 (0.9, 1.3) | 1.1 (0.9, 1.5) | 0.619 |
| **Medications** |  |  |  |  |
| Aspirin, n (%) | 46 (98) | 22 (96) | 24 (100) | 0.228 |
| P2Y<sub>12</sub> receptor inhibitor, n (%) | 46 (98) | 22 (96) | 24 (100) | 0.228 |
| β-blocker, n (%) | 37 (79) | 17 (74) | 20 (83) | 0.429 |
| ACE-i / ARB, n (%) | 31 (66) | 14 (61) | 17 (71) | 0.471 |
| Statin, n (%) | 46 (98) | 22 (96) | 24 (100) | 0.228 |
| Ezetimibe, n (%) | 14 (30) | 6 (26) | 8 (33) | 0.587 |
| **Sleep parameters** |  |  |  |  |
| Measurement time of pulse oximetry, min | 400±98 | 389±98 | 410±97 | 0.473 |
| Baseline mean 3% ODI, events/hour | 27.0±13.5 | 30.5±16.5 | 23.6±9.6 | 0.085 |
| Severe SDB (mean 3% ODI ≥30), n (%) | 14 (30) | 8 (35) | 6 (25) | 0.463 |
| Average SpO<sub>2</sub>, % | 93.4±1.5 | 93.1±1.8 | 93.7±1.2 | 0.173 |
| Minimum SpO<sub>2</sub>, % | 78.3±9.1 | 77.6±9.0 | 78.9±9.3 | 0.637 |
| Cumulative time with SpO<sub>2</sub> <90%, min | 15 (3, 65) | 22 (3, 79) | 14 (3, 46) | 0.463 |

ACE-i, angiotensin-converting enzyme inhibitor; ARB, angiotensin receptor blocker; CAD, coronary artery disease; CPAP, continuous positive airway pressure; Hs-CRP, high-sensitivity C-reactive protein; IMT, intima media thickness; NT-proBNP, N-terminal pro-brain natriuretic peptide. ODI, oxygen desaturation index; SDB, sleep-disordered breathing; SpO<sub>2</sub>, saturation of percutaneous oxygen.

### Procedural and imaging characteristics

Table 2 shows the procedural and imaging characteristics of the culprit and non-culprit lesions at baseline. Regarding the culprit lesion, 73 were treated using drug-eluting stents or drug-coated balloons. There were no significant differences in other angiographic data between the two groups except for lesion length. Regarding non-culprit lesions, 51 were measured using NIRS-IVUS. The non-culprit lesion totally had plaque burden 60±17%, total atheroma volume 193±158 mm^3^ and PAV 47.7±11.3%, including a plaque morphology of fibrous plaque 74%, soft plaque 15%, and calcified plaque 53%. There were no significant differences in these IVUS morphological data between the two groups.

**Table 2.** Procedural and imaging characteristics.

|  | Overall<br>(n=47) | CPAP<br>group (n=23) | Non-CPAP<br>group (n=24) | *p* value |
| --- | --- | --- | --- | --- |
| **Non-culprit lesion** |  |  |  |  |
| <u>Non-culprit lesion site</u> |  |  |  |  |
| Right coronary artery disease, n (%) | 14 (30) | 6 (26) | 8 (33) | 0.587 |
| Left anterior descending artery, n (%) | 17 (36) | 7 (30) | 10 (42) | 0.422 |
| Left circumflex artery, n (%) | 13 (28) | 8 (35) | 5 (21) | 0.234 |
| Left main trunk, n (%) | 7 (15) | 4 (17) | 3 (13) | 0.637 |
| <u>IVUS data</u> |  |  |  |  |
| Pull-back length, mm | 86.7±29.3 | 87.6±32.4 | 85.8±25.9 | 0.829 |
| Non-culprit lesion length, mm | 24.1±12.8 | 26.9±16.4 | 21.3±7.9 | 0.139 |
| Plaque morphology |  |  |  |  |
| Fibrous plaque, n (%) | 35 (74) | 19 (83) | 16 (67) | 0.207 |
| Soft plaque, n (%) | 7 (15) | 4 (17) | 3 (13) | 0.637 |
| Calcified plaque, n (%) | 25 (53) | 11 (48) | 14 (58) | 0.470 |
| Plaque burden at minimal lumen area, % | 60±17 | 63±15 | 57±19 | 0.193 |
| Vessel volume, mm<sup>3</sup> | 394±293 | 435±372 | 355±191 | 0.358 |
| Total atheroma volume, mm<sup>3</sup> | 193±158 | 207±197 | 179±109 | 0.545 |
| Percentage atheroma volume, % | 47.7±11.3 | 46.9±12.7 | 48.4±9.8 | 0.651 |
| <u>NIRS data</u> |  |  |  |  |
| Total lesion LCBI | 32 (0, 83) | 24 (1, 89) | 35 (0, 74) | 0.974 |
| MaxLCBI<sub>4mm</sub> | 111 (0, 260) | 111 (8, 279) | 123 (0, 214) | 0.629 |
| MaxLCBI<sub>10mm</sub> | 61 (0, 156) | 54 (2, 218) | 72 (0, 104) | 0.602 |
| Large LRP (maxLCBI<sub>4mm</sub> ≥400), n (%) | 5 (11) | 3 (13) | 2 (8) | 0.600 |
| **Culprit lesion** |  |  |  |  |
| <u>Culprit lesion site</u> |  |  |  |  |
| Right coronary artery disease, n (%) | 21 (45) | 10 (43) | 11 (46) | 0.871 |
| Left anterior descending artery, n (%) | 32 (68) | 16 (70) | 16 (67) | 0.831 |
| Left circumflex artery, n (%) | 19 (40) | 11 (48) | 8 (33) | 0.311 |
| Left main trunk, n (%) | 1 (2) | 0 (0) | 1 (4) | 0.322 |
| <u>Angiographic data</u> |  |  |  |  |
| Reference diameter, mm | 2.94±0.57 | 2.94±0.50 | 2.94±0.62 | 0.961 |
| Minimal luminal diameter, mm | 0.79±0.45 | 0.87±0.46 | 0.70±0.46 | 0.213 |
| Culprit lesion length, mm | 31.1±12.9 | 26.7±9.5 | 35.3±15.5 | 0.027 |
| Multi-vessel disease, n (%) | 29 (62) | 14 (61) | 15 (63) | 0.909 |
| <u>Procedural devices</u> |  |  |  |  |
| Drug-eluting stent, n (%) | 46 (98) | 23 (100) | 23 (96) | 0.243 |
| Stent size, mm | 2.89±0.48 | 2.82±0.39 | 2.96±0.55 | 0.317 |
| Stent length, mm | 32.3±13.3 | 27.7±9.5 | 37.0±16.2 | 0.021 |

CPAP, continuous positive airway pressure; IVUS, intravascular ultrasound; LCBI, lipid-core burden index; LRP, lipid-rich plaque; MaxLCBI<sub>4mm</sub>, maximum lipid core burden index calculated for every 4-mm segment; MaxLCBI<sub>10mm</sub>, maximum lipid core burden index calculated for every 10-mm segment; NIRS, near infrared spectroscopy.

The median total lesion LCBI, maxLCBI_4mm_, and maxLCBI_10mm_ were 32 (IQR, 0-83), 111 (IQR, 0-260), and 61 (IQR, 0-156), respectively, and a large lipid-rich plaque accounted for 11%. However, there was no significant difference in these LCBI values between the two groups. In addition, the correlations between PAV and maxLCBI_4mm_ and maxLCBI_10mm_ were significantly positive (r=0.33, *p*=0.023; and r=0.35, *p*=0.018, respectively).

### Sleep assessment and CPAP adherence

Table 3 shows CPAP adherence and sleepiness. Among the CPAP group, the median daily use of CPAP device and median monthly use of CPAP device were 4.3 (IQR, 2.6-5.2) hour/night and 27 (IQR, 22-29) day/month, respectively. In addition, adequate CPAP treatment were observed in 12 patients (52%).

**Table 3.**
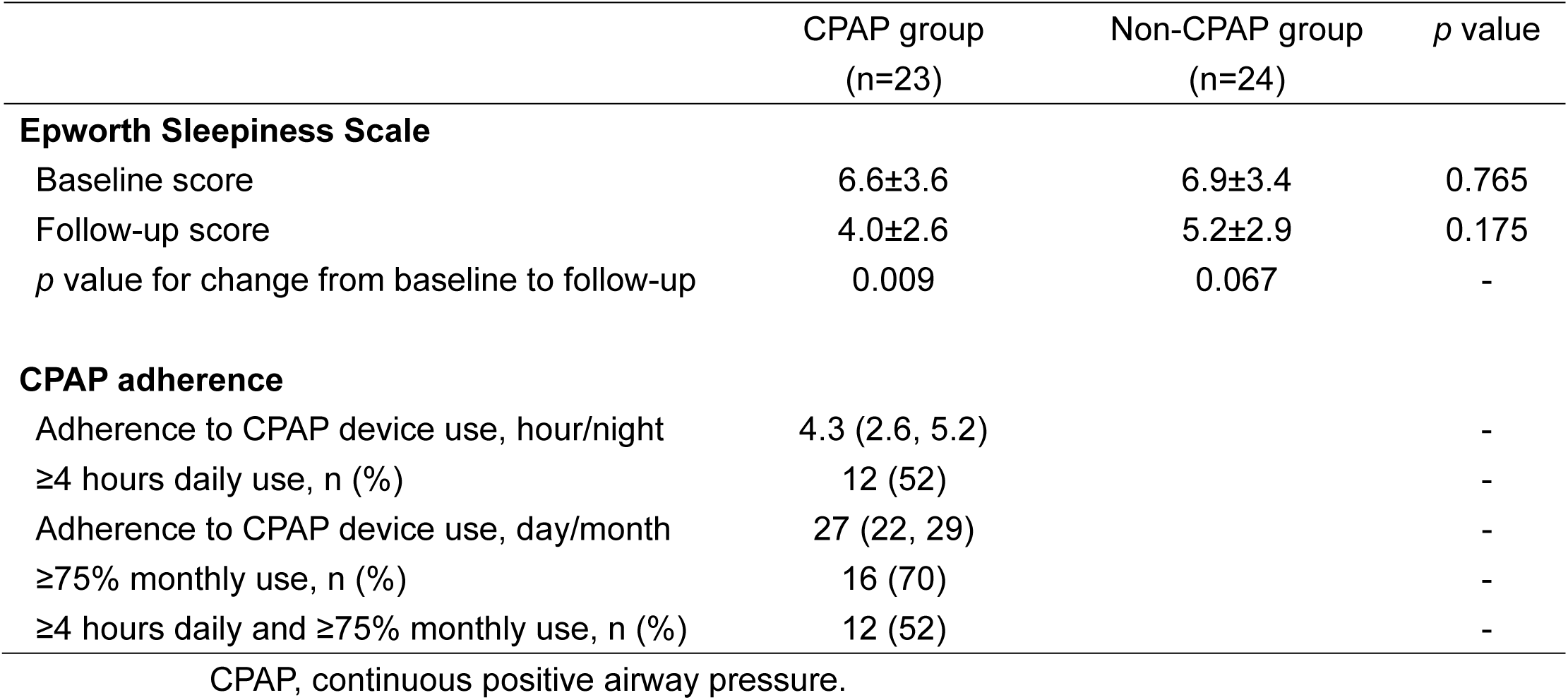
Sleep assessment and CPAP adherence.

|  | CPAP group<br>(n=23) | Non-CPAP group<br>(n=24) | *p* value |
| --- | --- | --- | --- |
| **Epworth Sleepiness Scale** |  |  |  |
| Baseline score | 6.6±3.6 | 6.9±3.4 | 0.765 |
| Follow-up score | 4.0±2.6 | 5.2±2.9 | 0.175 |
| *p* value for change from baseline to follow-up | 0.009 | 0.067 | - |
| **CPAP adherence** |  |  |  |
| Adherence to CPAP device use, hour/night | 4.3 (2.6, 5.2) |  | - |
| ≥4 hours daily use, n (%) | 12 (52) |  | - |
| Adherence to CPAP device use, day/month | 27 (22, 29) |  | - |
| ≥75% monthly use, n (%) | 16 (70) |  | - |
| ≥4 hours daily and ≥75% monthly use, n (%) | 12 (52) |  | - |

CPAP, continuous positive airway pressure.

There was a significant difference in the average Epworth Sleepiness Scale score between baseline and follow-up among the 23 patients who received CPAP treatment (baseline Epworth Sleepiness Scale score vs. follow-up Epworth Sleepiness Scale score, 6.6±3.6 vs. 4.0±2.6, *p*=0.009), but not among the 24 patients who did not receive CPAP treatment.

### Impact of CPAP on coronary plaques and cardiovascular events

Representative image of the coronary plaque detected by NIRS-IVUS and non-culprit lesion site revealed by angiography in patient on CPAP are shown in Figure 2. Table 4 presents the results for the primary and secondary endpoints. The absolute change in PAV defined as primary endpoint was −1.90±2.63% in the CPAP group and 0.82±4.67% in the non-CPAP group, respectively, as shown in Figure 3(A). The adjusted difference of PAV in the two groups was −2.81±1.10% by analysis of covariance (*p*=0.014). With regard to the secondary endpoints, the CPAP group had a significantly higher absolute change in total atheroma volume compared with the non-CPAP group (−20±35 mm^3^ vs. −2±21 mm^3^, *p*=0.041). However, there were no significant differences in absolute changes in maxLCBI_4mm_, maxLCBI_10mm_, maximum intima-media thickness, or high-sensitivity C-reactive protein levels, and no deaths occurred during the follow-up period. Interestingly, the CPAP group had a significantly lower incidence rate of MACCE than that in the non-CPAP group, mainly driven by a decrease in target vessel revascularization (4% vs. 25%, *p*=0.037).

**Figure 2.**
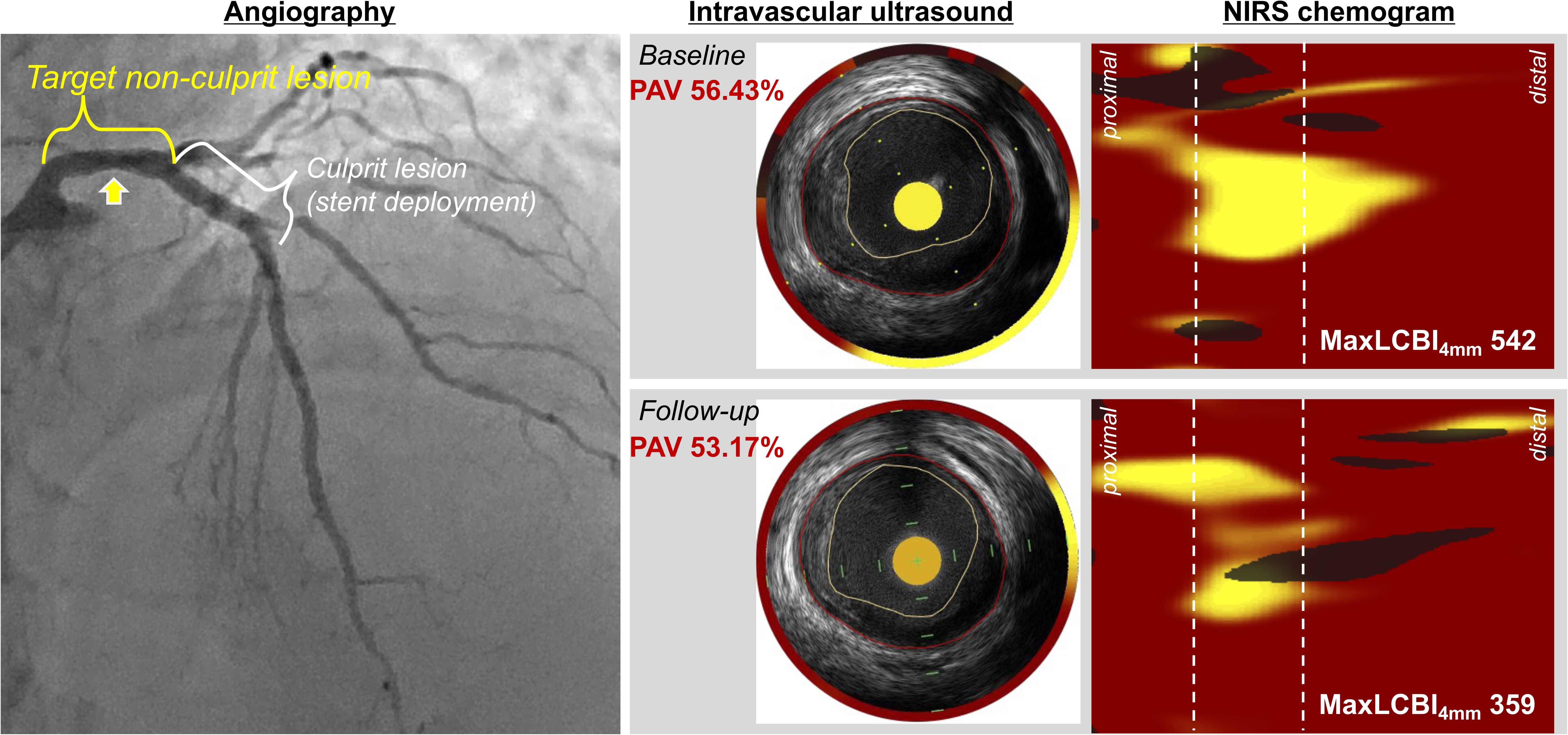
Representative image of patient with CPAP treatment. The left panel shows the angiographic image, including the non-culprit lesion site (yellow curly bracket) and culprit lesion site (white curly bracket); the right upper panel and lower panel show the IVUS images and NIRS chemogram at baseline and follow-up, respectively, in a patient who received CPAP treatment. The IVUS images show the non-culprit lesion site indicated by a yellow arrow on the angiographic image, and the PAV improved from 56.43% at baseline to 53.17% at follow-up. In addition, the NIRS chemogram shows an improvement in maxLCBI_4mm_ from 542 to 359. CPAP, continuous positive airway pressure; IVUS, intravascular ultrasound; MaxLCBI_4mm_, maximum lipid core burden index calculated for every 4-mm segment; NIRS, near-infrared spectroscopy; PAV, percentage atheroma volume.

**Figure 3.**
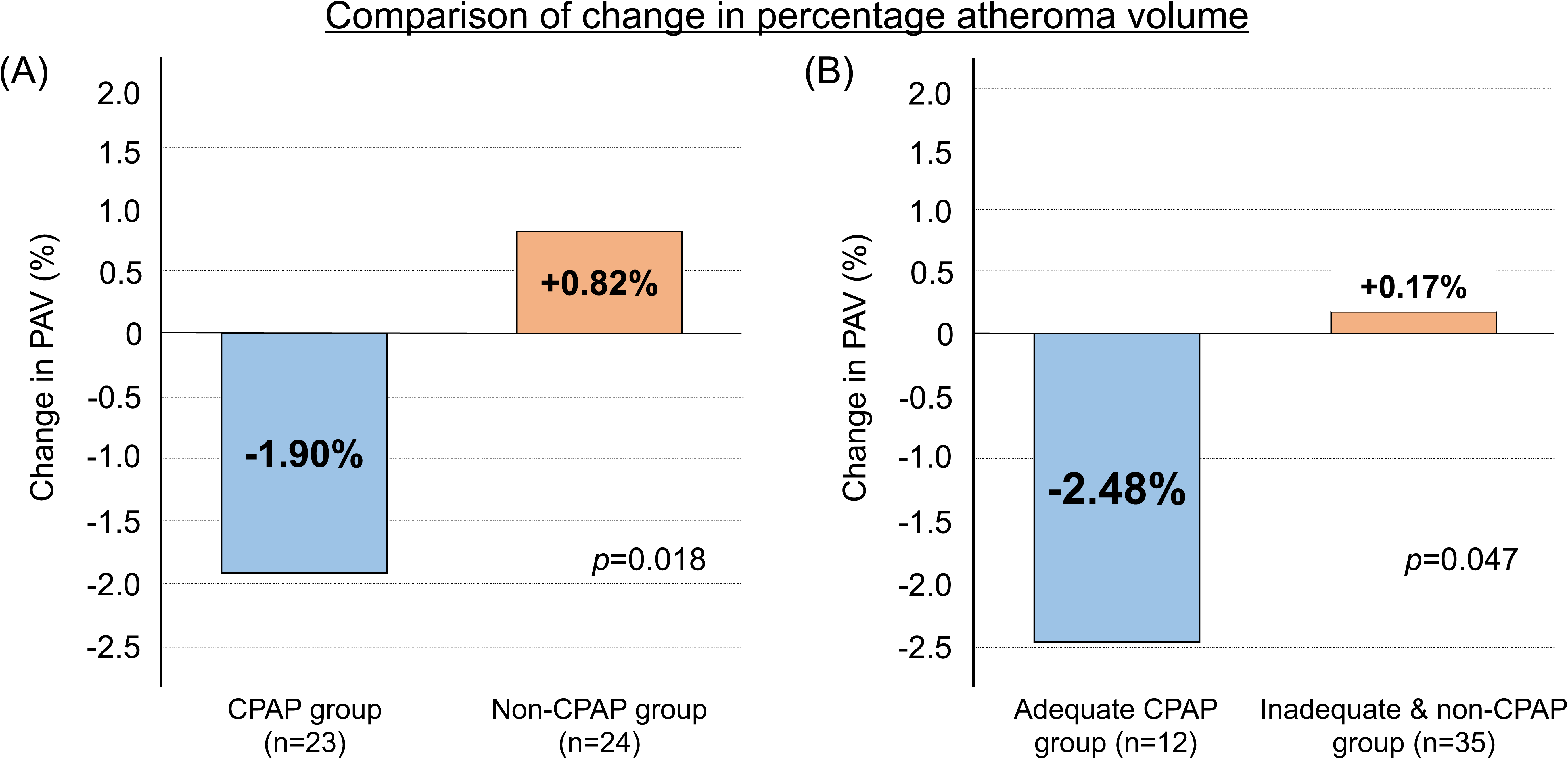
Comparison of change in percentage atheroma volume. (A) Comparison between the CPAP group and non-CPAP group (B) Comparison between the adequate CPAP group, and inadequate and non-CPAP group CPAP, continuous positive airway pressure; PAV, percentage atheroma volume.

**Table 4.**
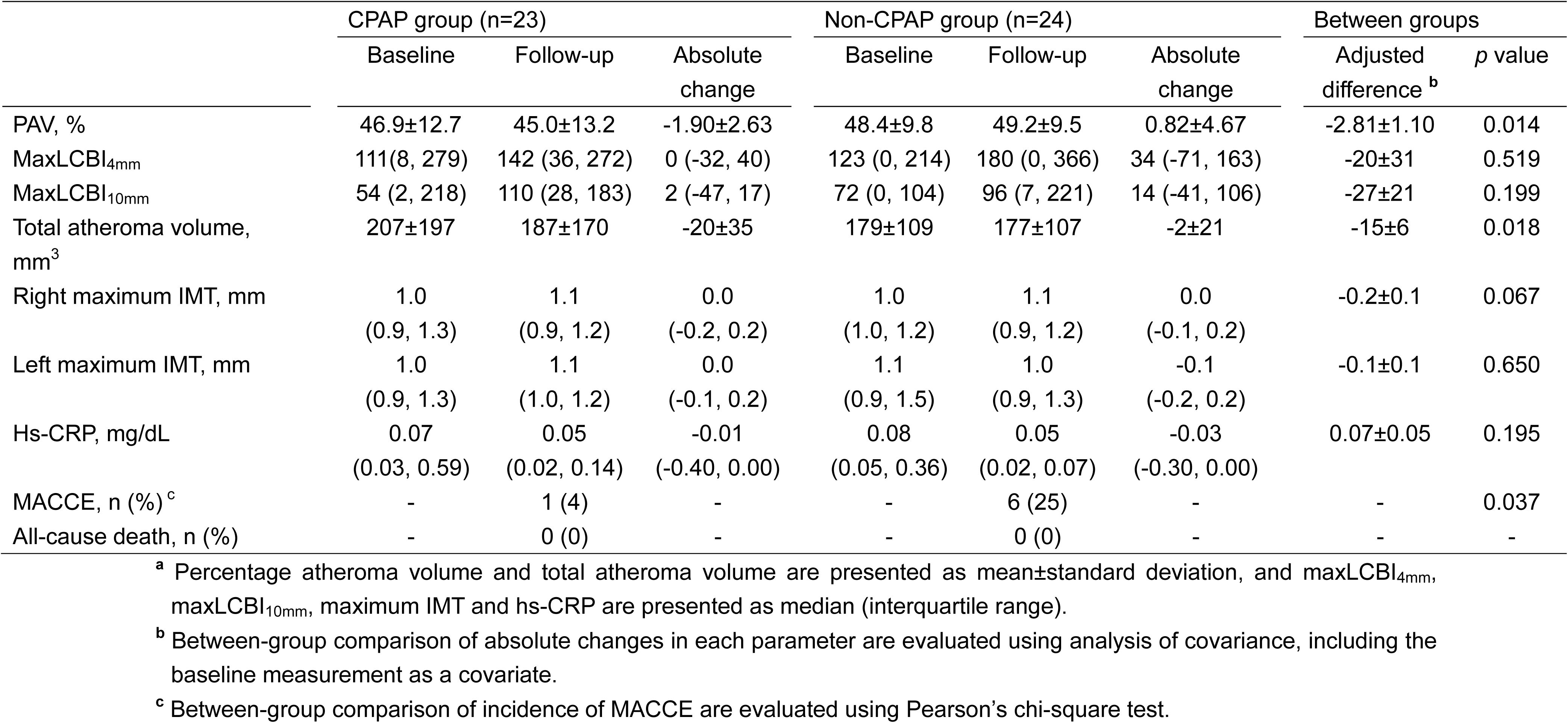
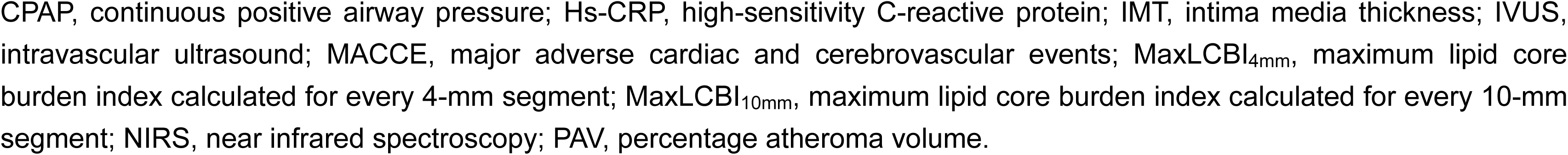
Primary and secondary endpoints^a^.

|  | CPAP group (n=23) |  |  | Non-CPAP group (n=24) |  |  | Between groups |  |
| --- | --- | --- | --- | --- | --- | --- | --- | --- |
|  | Baseline | Follow-up | Absolute change | Baseline | Follow-up | Absolute change | Adjusted difference <sup>b</sup> | p value |
| PAV, % | 46.9±12.7 | 45.0±13.2 | -1.90±2.63 | 48.4±9.8 | 49.2±9.5 | 0.82±4.67 | -2.81±1.10 | 0.014 |
| MaxLCBI<sub>4mm</sub> | 111(8, 279) | 142 (36, 272) | 0 (-32, 40) | 123 (0, 214) | 180 (0, 366) | 34 (-71, 163) | -20±31 | 0.519 |
| MaxLCBI<sub>10mm</sub> | 54 (2, 218) | 110 (28, 183) | 2 (-47, 17) | 72 (0, 104) | 96 (7, 221) | 14 (-41, 106) | -27±21 | 0.199 |
| Total atheroma volume, mm<sup>3</sup> | 207±197 | 187±170 | -20±35 | 179±109 | 177±107 | -2±21 | -15±6 | 0.018 |
| Right maximum IMT, mm | 1.0<br>(0.9, 1.3) | 1.1<br>(0.9, 1.2) | 0.0<br>(-0.2, 0.2) | 1.0<br>(1.0, 1.2) | 1.1<br>(0.9, 1.2) | 0.0<br>(-0.1, 0.2) | -0.2±0.1 | 0.067 |
| Left maximum IMT, mm | 1.0<br>(0.9, 1.3) | 1.1<br>(1.0, 1.2) | 0.0<br>(-0.1, 0.2) | 1.1<br>(0.9, 1.5) | 1.0<br>(0.9, 1.3) | -0.1<br>(-0.2, 0.2) | -0.1±0.1 | 0.650 |
| Hs-CRP, mg/dL | 0.07<br>(0.03, 0.59) | 0.05<br>(0.02, 0.14) | -0.01<br>(-0.40, 0.00) | 0.08<br>(0.05, 0.36) | 0.05<br>(0.02, 0.07) | -0.03<br>(-0.30, 0.00) | 0.07±0.05 | 0.195 |
| MACCE, n (%) <sup>c</sup> | - | 1 (4) | - | - | 6 (25) | - | - | 0.037 |
| All-cause death, n (%) | - | 0 (0) | - | - | 0 (0) | - | - | - |

<sup>a</sup> Percentage atheroma volume and total atheroma volume are presented as mean±standard deviation, and maxLCBI<sub>4mm</sub>, maxLCBI<sub>10mm</sub>, maximum IMT and hs-CRP are presented as median (interquartile range).
<sup>b</sup> Between-group comparison of absolute changes in each parameter are evaluated using analysis of covariance, including the baseline measurement as a covariate.
<sup>c</sup> Between-group comparison of incidence of MACCE are evaluated using Pearson's chi-square test.
CPAP, continuous positive airway pressure; Hs-CRP, high-sensitivity C-reactive protein; IMT, intima media thickness; IVUS, intravascular ultrasound; MACCE, major adverse cardiac and cerebrovascular events; MaxLCBI<sub>4mm</sub>, maximum lipid core burden index calculated for every 4-mm segment; MaxLCBI<sub>10mm</sub>, maximum lipid core burden index calculated for every 10-mm segment; NIRS, near infrared spectroscopy; PAV, percentage atheroma volume.

When only patients with adequate CPAP treatment were included in the analysis, the absolute change in PAV was −2.48±2.44% and that in maxLCBI_4mm_ was 13 (−3, 37). The atheroma volume was significantly lower in the adequate CPAP group then in the inadequate and non-CPAP groups, as shown in Figure 3(B), but not the extent of lipid-rich plaque represented by maxLCBI_4mm_ (data not shown).

## Discussion

The major findings of this study were: 1) this study is the first report on the effective treatment of SDB with CPAP for 12-months on coronary plaque volume and lipid-rich plaque analyzed by NIRS-IVUS in patients with stable coronary artery disease; 2) CPAP treatment for SDB-induced coronary plaque regression on IVUS with no significant change in the extent of lipid-rich plaque on NIRS; 3) patients with CPAP treatment had significantly lower cumulative clinical events compared with patients with no CPAP treatment; and 4) a sub-analysis showed that adequate CPAP treatment had a greater effect on plaque regression than inadequate and non-CPAP treatment.

The mechanism of vascular damage in patients with SDB involves oxidative stress, systemic inflammation, sympathetic activation, and then endothelial dysfunction, caused by the main acute consequences of SDB, such as intermittent hypoxia.^23^ This arteriosclerotic state leads to the development of coronary plaques in patients with SDB, and the degree of SDB is associated with the burden and vulnerability of non-calcified plaque detected using IVUS.^6^ Thus, it should be discussed how these causal relationships are affected by CPAP treatment. The reduction in inflammation and improvement of endothelial function in the coronary arteries are associated with a proportional reduction in the atherosclerosis and cardiovascular events.^24^ It has recently been reported that the assessment of atherosclerotic carotid and aortic plaque activity using fluorodeoxyglucose is useful in detecting the accumulation of cholesterol-engorged macrophages, and CPAP treatment for patients with SDB contributes to an average reduction in fluorodeoxyglucose uptake.^25^ In addition, a retrospective observational study on the optical coherence tomography findings in patients with SDB with stable coronary artery disease showed that the CPAP-SDB group had a significantly lower macrophage grading than the untreated-SDB group.^26^ CPAP suppresses inflammatory macrophages and renders coronary plaques inactive.^27^ In this study, there was no significant difference in the absolute change in high-sensitivity C-reactive protein levels between the groups; however, CPAP treatment may improve the inflammatory response from a microscopic perspective. Also, the changes from baseline to follow-up in LDL-C levels, hemoglobin A1c levels and insulin resistance promoted endothelial dysfunction were not significantly different with CPAP treatment in this study. However, the previous studies have shown that CPAP treatment contributes to the improvement and normalization of endothelial function; thus, CPAP treatment may have an effect on plaque regression due to the improvement of endothelial function.^28, 29^

Furthermore, among the many reports on the clinical effects of coronary plaque regression and stability, the most recommended therapy is LDL-C lowering agents, such as statins, ezetimibe, and proprotein convertase subtilisin kenin 9 inhibitor.^22, 30, 31^ A previous study showed that the statin-alone therapy and dual LDL-C lowering therapy with statins and ezetimibe contributed in patients with stable coronary artery disease for the plaque regression represented as decline in PAV (−0.7% and −1.2%, respectively).^22^ In this study, setting a 2.20% difference in the primary endpoint was used, and a sample size of 100 patients was planned when the protocol was created.^10^ As a result, only approximately half of them were included in the final analysis, because the case recruitment was difficult due to the pandemic of COVID-19. However, it seems that a significant difference was recognized with a between-group difference of −2.81%, which exceeds expectations. Therefore, it is reasonable to conclude that CPAP leads to coronary plaque regression in patients with SDB.

By contrast, CPAP is expected to contribute to plaque stability by preventing atherosclerosis. However, this study found no significant difference in the extent of lipid-rich plaques between the two groups. As patients with stable coronary artery disease tend to have fewer lipid-rich plaques than those with acute coronary syndrome, it is conceivable that the absolute change in the maxLCBI_4mm_ value is unlikely to result in a significant difference.^17, 18^ In this study, the proportion of large lipid-rich plaque was five patients (11%), and the use of CPAP in patients with large lipid-rich plaques improved the absolute change in the maxLCBI_4mm_ value compared with the non-use of CPAP (−93 vs. −37), whereas the use of CPAP in patients without large lipid-rich plaques did not improve the absolute change in the maxLCBI_4mm_ value compared with the non-use of CPAP (+22 vs. +36). Since this study only targets patients with stable coronary artery disease, maxLCBI_4mm_ value is relatively low, and the effects of CPAP on plaque vulnerability assessed by NIRS-IVUS should be verified for abundant lipid core. Thus, the further investigations targeting patients with acute coronary syndrome who have greater lipid-rich plaques than those with stable coronary artery disease are necessary.

Furthermore, this study showed that the CPAP group had a significantly lower incidence rate of MACCE compared with the non-CPAP group, mainly due to a decrease in target vessel revascularization. However, the randomized clinical trials targeting patients diagnosed with obstructive sleep apnea and any cardiovascular diseases showed that the all-cause death and adverse cardiovascular events were not significantly reduced in patients received CPAP treatment.^32^ The poor adherence to CPAP treatment, shown as the average use time of ≤4 hours/night, is an important limitation as this causal relationship.^33^ In this study, good adherence to CPAP treatment may contribute to the prevention of atherosclerosis at de-novo and in-stent sites in patients with SDB who received CPAP treatment.

This study has several limitations that require consideration. First, as this was a prospective, randomized, open-label, single-center study, unknown confounding factors might have affected the study outcomes regardless of analytical adjustments, and the relatively small number of enrolled patients may have limited the statistical power of the study. Second, this study had half the original planned sample size because of difficulties in case recruitment caused by the pandemic of COVID-19. However, the expected plaque regression effect was greater and a significant difference was observed between the two groups. Third, pulse oximeter inherently underestimates respiratory status during sleep compared to polysomnography. However, some reports showed that patients who diagnosed with a diagnosis of SDB based on the results of pulse oximeter and underwent PCI involved in worse clinical outcomes and had larger and more unstable plaques.^4, 6^ Therefore, this study was considered the validity of previous research and the feasibility in clinical practice, and CPAP was also introduced in the evaluation in the evaluation by pulse oximeter.

## Conclusions

To our knowledge, this study is the first to report on the effect of CPAP treatment on coronary plaque volume and lipid-rich plaques in patients with SDB with stable coronary artery disease who underwent PCI, using NIRS-IVUS. CPAP treatment contributed to a decline in coronary plaque volume, and the prevention of cardiovascular events, mainly driven by a decrease in target vessel revascularization but not by the mitigation of lipid-rich plaques. In particular, adequate CPAP treatment had a greater effect on coronary plaque regression.

## Data Availability

The datasets during and/or analyzed during the current study are available from the corresponding author with reasonable request.

## Abbreviations list

CPAP: continuous positive airway pressure
IVUS: intravascular sound
LCBI: lipid core burden index
LDL-C: low-density lipoprotein cholesterol
MACCE: major adverse cardiac and cerebrovascular events
MaxLCBI_4mm/10mm_: maximum lipid core burden index calculated for every 4-mm / 10-mm segment
NIRS: near-infrared spectroscopy
PAV: percentage atheroma volume
PCI: percutaneous coronary intervention
SDB: sleep-disordered breathing

## Acknowledgments

The authors are grateful to the staff of the Department of Cardiovascular Medicine at Juntendo University. The authors also appreciate the secretarial assistance of Yumi Nozawa, and the clinical trial coordinator of Yumiko Kanehiro.

